# Nowcasting and forecasting provincial-level SARS-CoV-2 case positivity using google search data in South Africa

**DOI:** 10.1101/2020.11.04.20226092

**Authors:** Elaine O. Nsoesie, Karla Therese L. Sy, Olubusola Oladeji, Raesetje Sefala, Brooke E. Nichols

## Abstract

Data from non-traditional data sources, such as social media, search engines, and remote sensing, have previously demonstrated utility for disease surveillance. Few studies, however, have focused on countries in Africa, particularly during the SARS-CoV-2 pandemic. In this study, we use searches of COVID-19 symptoms, questions, and at-home remedies submitted to Google to model COVID-19 in South Africa, and assess how well the Google search data forecast short-term COVID-19 trends. Our findings suggest that information seeking trends on COVID-19 could guide models for anticipating COVID-19 trends and coordinating appropriate response measures.

## INTRODUCTION

The SARS-CoV-2 pandemic has infected millions of people globally, and continues to spread rapidly in many countries. During the initial COVID-19 outbreaks in March 2020, countries in Africa were able to contain large waves of infections with the early implementation of containment measures; however, as lockdowns and restrictions have eased and testing has expanded, cases in Africa have steadily increased even with the scale up of national and provincial responses.^1^ As of August 2020, more than half of Africa’s cases were recorded in South Africa, despite only comprising 5% of the continent’s total population - largely due to South Africa’s rapid expansion of Polymerase Chain Reaction (PCR) testing capacity for SARS-CoV-2 diagnosis.^2,3^ After the first wave, which occurred in the South African winter, seroprevalence survey estimates that between 29%-40% have been infected, varying strongly by geography.^4,5^ Moreover, as of October 2020, cases have been steadily increasing in South Africa.^6,7^ A number of factors facilitate increased COVID-19 transmission in Africa, including weak health care systems,^8^ high prevalence of other diseases such as HIV and TB,^9-12^ and the inability to adhere to physical distancing and other intervention strategies.^13^

Although the current reported mortality burden is lower than had been projected by public health experts, the pandemic has highlighted gaps in surveillance systems.^14-16^ Surveillance systems could incorporate non-traditional sources of information that can provide insights into local spread of disease. Data from non-traditional data sources, such as social media, search engines, and remote sensing have demonstrated usefulness for disease surveillance. While there are indications that these data could be useful for supplementing traditional surveillance systems, few studies have focused on countries in Africa.^17,18^ Specifically, searches of disease-related terms (such as, COVID-19) could reveal trends in cases. A recent report estimated that 56.2% of the South African population had access to the internet in 2017.^19^ Data submitted on search engines could assist in monitoring COVID-19 trends and response to interventions, which would assist in mitigating the spread of COVID-19 in South Africa, and to trigger rapid resource allocation.

In this paper, we used searches of COVID-19 symptoms, questions, and at-home remedies submitted to Google to model expected current and future COVID-19 cases in South Africa. We assessed how well the search data forecasted near future COVID-19 trends, information vital for the coordination of rapid response measures.

## METHODS

### COVID-19 Case Data

We collected data on reported COVID-19 cases and number of tests administered from the South African National Institute of Communicable Diseases from April 19th 2020 to August 29th, 2020.^20^ The National Department of Health defined a COVID-19 case as laboratory confirmation of COVID-19 disease based on PCR tests. Confirmed COVID-19 case counts and tests were reported from all nine provinces in South Africa on a daily basis. Since variability in testing during the pandemic can affect the number of cases detected, the main outcome of the study was weekly test positivity rate, which is the number of positive cases divided by the number of individuals tested. This metric has previously been used to gauge the current severity of the epidemic.^21^

### Google Search Data

We made a list of forty-three Google Search terms crowdsourced from infectious disease modelers and data scientists in South Africa (Table 1). The term list included questions about COVID-19, symptoms, and at-home remedies used in South Africa. We downloaded weekly search volume from the Google API for each of the phrases in the list for all provinces (i.e., Gauteng, Eastern Cape, Free State, KwaZulu-Natal, Limpopo, Mpumalanga, North West, Northern Cape, and Western Cape). The search volume for each of the terms relative to the search activity in the region is normalized by Google.

**Table 1:** List of search terms

| Number | Categories | Search Terms |
| --- | --- | --- |
| 1.) | Questions about COVID-19 symptoms | what are the symptoms of COVID-19, flu symptoms, symptoms of COVID-19, coronavirus symptoms, COVID symptoms, what are the symptoms of COVID |
| 2.) | Disease symptoms | Cough, fever, flu, sore throat, headache, runny nose, shortness of breath, nausea, fatigue, chill, vomiting, loss of appetite, chest pain, loss of taste, loss of smell, chest pressure, difficulty breathing, muscle ache, diarrhea, congestion |
| 3.) | COVID-19 terms and questions | coronavirus, COVID, coro, COVID-19, corona viris, how do I know I have COVID, what happens if I get COVID, can I get COVID |
| 4.) | At-home remedies | Ginger tea, zinc, zimplex, eucalyptus oil, COVID supplements, COVID home remedy, umhlonyane, Lengana, calceterol |

### Statistical Analysis

We first calculated the association between the search terms and COVID-19 case data using Pearson correlation to assess how well searches related to the test positivity rate. Next, we fit several multivariable regression models using Elastic Net, a regularization and variable selection method, previously used and validated for modeling influenza-like illness (ILI) trends.^22-24^ We selected the Elastic Net modeling approach because it has many advantages, including good performance when the number of observations are small and the number of covariates are arge.^22,25^ The multivariable regression model is defined as, 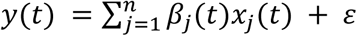where x represents search terms for each of the provinces, y is the case positivity rate, *β* are the model estimated coefficients and *t* is time. To determine the best model for each province, we varied the value of *α* in the estimator given by 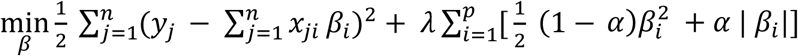 When *α* equals zero, the estimator equates to Ridge regression and Least Absolute Shrinkage and Selection Operator (LASSO) when *α* is one. We fitted separate models for each province and used the adjusted coefficient of determination and Root Mean Squared Error (RMSE) to assess model fit and forecast, as well as Pearson correlation to quantify their associations.

## RESULTS

### Associations between searches and COVID-19 cases

The two search terms with the most significant correlation with COVID-19 case positivity rate for each of the nine provinces were as follows (Figure 1): Western Cape (COVID symptoms [r=0.878] and shortness of breath [0.729]), Eastern Cape (eucalyptus oil [0.926] and COVID symptoms [0.858]), Northern Cape (eucalyptus oil [0.665] and shortness of breath [0.588]), Free State (zinc [0.782] and COVID symptoms [0.890]), KwaZulu Natal (loss of smell [0.919] and COVID symptoms [0.882]), North West (COVID symptoms [0.794] and symptoms of COVID-19 [0.768]), Gauteng (loss of smell [0.899] and loss of taste [0.885]), Mpumalanga (COVID symptoms [0.762] and eucalyptus oil [0.653]), and Limpopo (loss of smell [0.719] and runny nose [0.673]). The search terms COVID symptoms, shortness of breath, eucalyptus oil, zinc, and loss of smell were repeated for multiple provinces.

**Figure 1:**
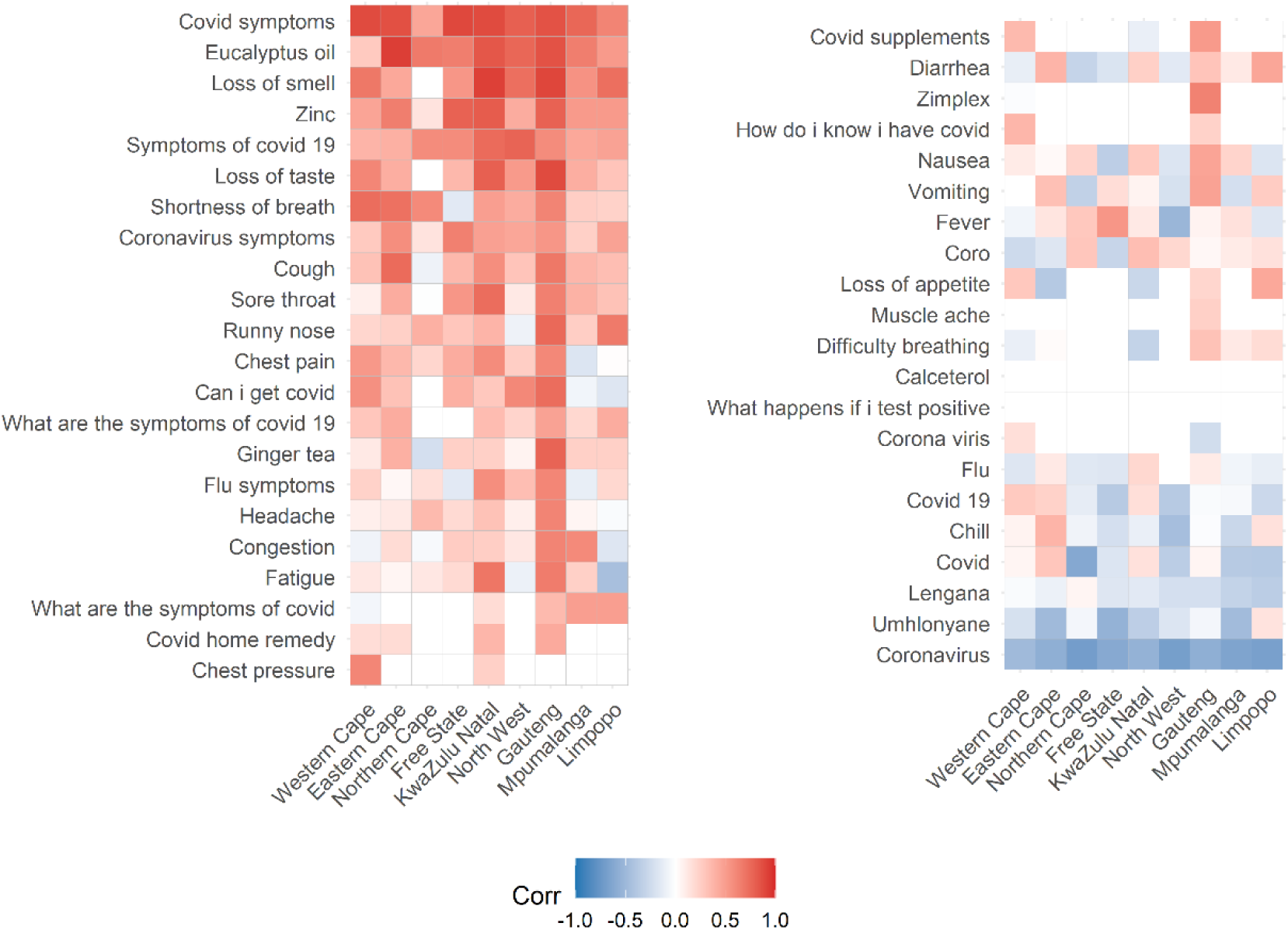
Correlation between search terms and COVID-19 case positivity rate. The correlations were significantly higher for some provinces (such as, Gauteng and Eastern Cape) compared to others especially for at-home remedies (e.g., Eucalyptus oil) and symptoms (e.g., loss of smell).

### Nowcasting and Forecasting COVID-19 cases

Elastic Net with α equal to one (i.e., LASSO regression) had the best model fit. The correlation between the model estimates and proportion of positive cases were 0.854, 0.935, 0.949, 0.912, 0.962, 0.871, 0.776, 0.798 and 0.992 for Limpopo, Mpumalanga, Gauteng, North West, KwaZulu Natal, Free State, Northern Cape, Western Cape and Eastern Cape, respectively (Figure 2, SI Figure 1). The corresponding root mean squared errors (RMSE) were 0.048, 0.045, 0.038, 0.056, 0.034, 0.068, 0.073, 0.044 and 0.015. Moreover, there was a significant drop and increase in reported Google searches during the first week and last weeks of August, respectively, which impacted estimates for several provinces. When search trends from the previous week were used to forecast cases for the next week, the models estimates of COVID-19 cases improved moderately (Figure 3).

**Figure 2:**
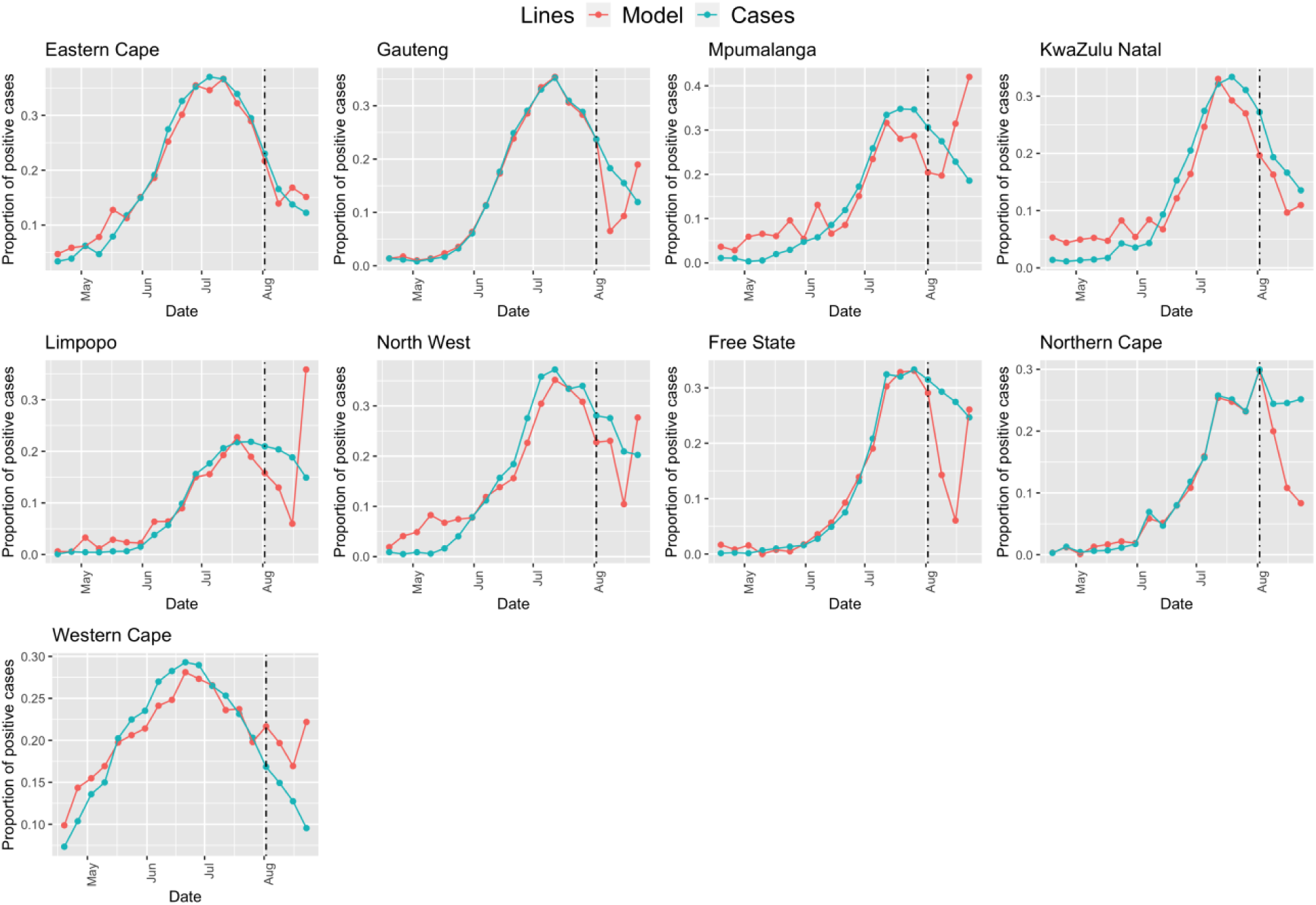
We fitted the model using data from April 19th 2020 to August 1st (indicated by the black vertical line), and for the remaining period, estimated proportion of positive COVID-19 cases for timepoint t, given search data at point t.

**Figure 3:**
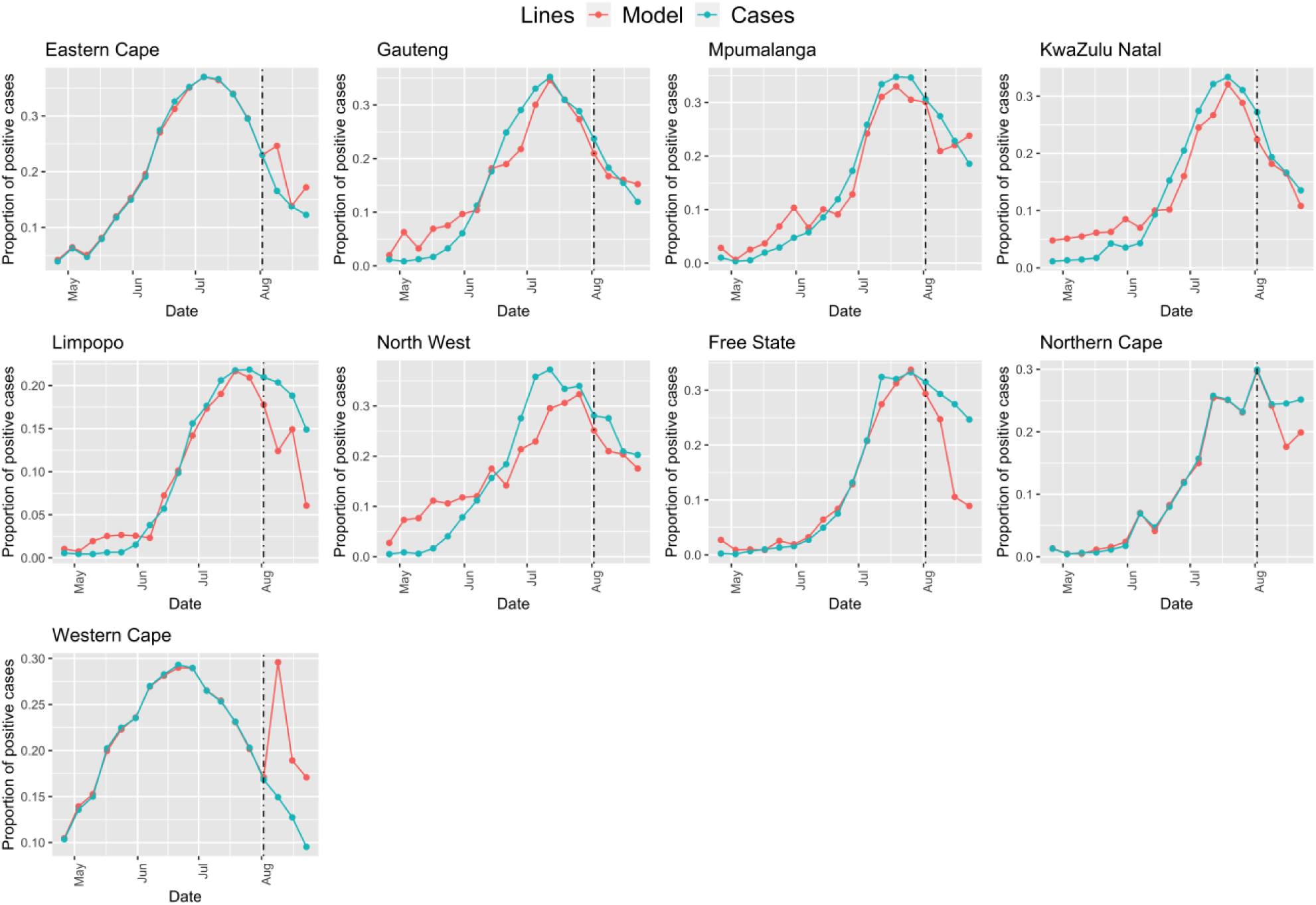
We fitted the model using data from April 19th 2020 to August 1st (indicated by the black vertical line), and from thereafter estimated proportion of positive COVID-19 cases for timepoint t+1, given search data at point t.

## DISCUSSION

Our work demonstrates that a number of Google Search terms were strongly correlated with test positivity rate, and this varied by province. Furthermore, our province-level models predicted COVID-19 test positivity with good model fit. This demonstrates the importance of geographically specific models for using Google search term data to predict future cases. The nowcast models suggest that people were already seeking information on remedies prior to getting tested in some provinces, such as KwaZulu Natal, since our results show that Google term searches peaked a week earlier than SARS-CoV-2 cases. This is consistent with the delay in onset of symptoms, seeking a test, and getting a test result. As such, our forecast models demonstrate that search data could be used to forecast short term trends in COVID-19 cases, as individuals likely seek information about symptoms and remedies prior to getting tested, during the first few days of symptom onset. Our findings suggest that information seeking trends on COVID-19 could guide models for anticipating COVID-19 trends, informing decision making and rapid resource allocation in South Africa. The use of these provincial-level google search terms can also be an early warning system that could trigger additional mitigation measures ahead of a rise in cases.

This study is important for several reasons. First, there is a paucity of studies demonstrating the usability of data from Internet sources for disease surveillance in sub-Saharan Africa. Second, we note that in addition to symptoms, at-home remedies used in managing COVID-19 symptoms were significant in estimating current and future COVID-19 case trends. This reinforces the need for developing contextualized approaches that take into account cultural and social practices.^18^ Lastly, geographical variability in our estimates suggest that factors as Internet usage, representation and socioeconomic factors should be considered when using digital data for disease surveillance.

There are some limitations to our study. The study period is short, therefore limiting the available data. However, even with the data limitations, we were able to create models with good fit; thus, as the SARS-CoV-2 pandemic continues, we would expect that additional data points would only improve model estimates. Due to the small sample size, confidence intervals (CI) were unreliable. However, CI can be estimated by adding an additional modeling step such as, a generalized linear model (see SI Figure 1 and 2). Second, there might be variability in the at-home remedies that were used in different provinces, and were not captured in our data. We aimed to include an inclusive and comprehensive list of appropriate search terms, and the input by several infectious disease scientists familiar with the South African context provided insight on province-specific cultural and social practices. The inclusion of any further provincial-specific at-home remedies in our search terms would only further improve the model fit. Third, if there are large changes to testing strategies used over time, that may affect the use of these models to forecast case-positivity. Should changes to the testing strategy occur, such as widespread use of rapid antigen diagnostics, or community testing, different outcome metrics may become more reliable for forecasting.

Despite these limitations, we demonstrate that there is value in using non-traditional data sources for disease surveillance in sub-Saharan Africa. As more people use the Internet for seeking and sharing health information, there will be more opportunities for developing digital surveillance tools that could supplement traditional surveillance systems. Given that COVID-19 cases in the African continent are increasing again, novel disease surveillance tools are invaluable for the coordination of rapid response measures.

## Data Availability

National Institute for Communicable Diseases COVID-19 testing and case data in South Africa are publicly available (link: https://www.nicd.ac.za). South Africa Google Trend data is available openly (link: https://trends.google.com/trends/?geo=ZA).

https://trends.google.com/trends/?geo=ZA

## SUPPLEMENTARY MATERIAL

**SI Figure 1:**
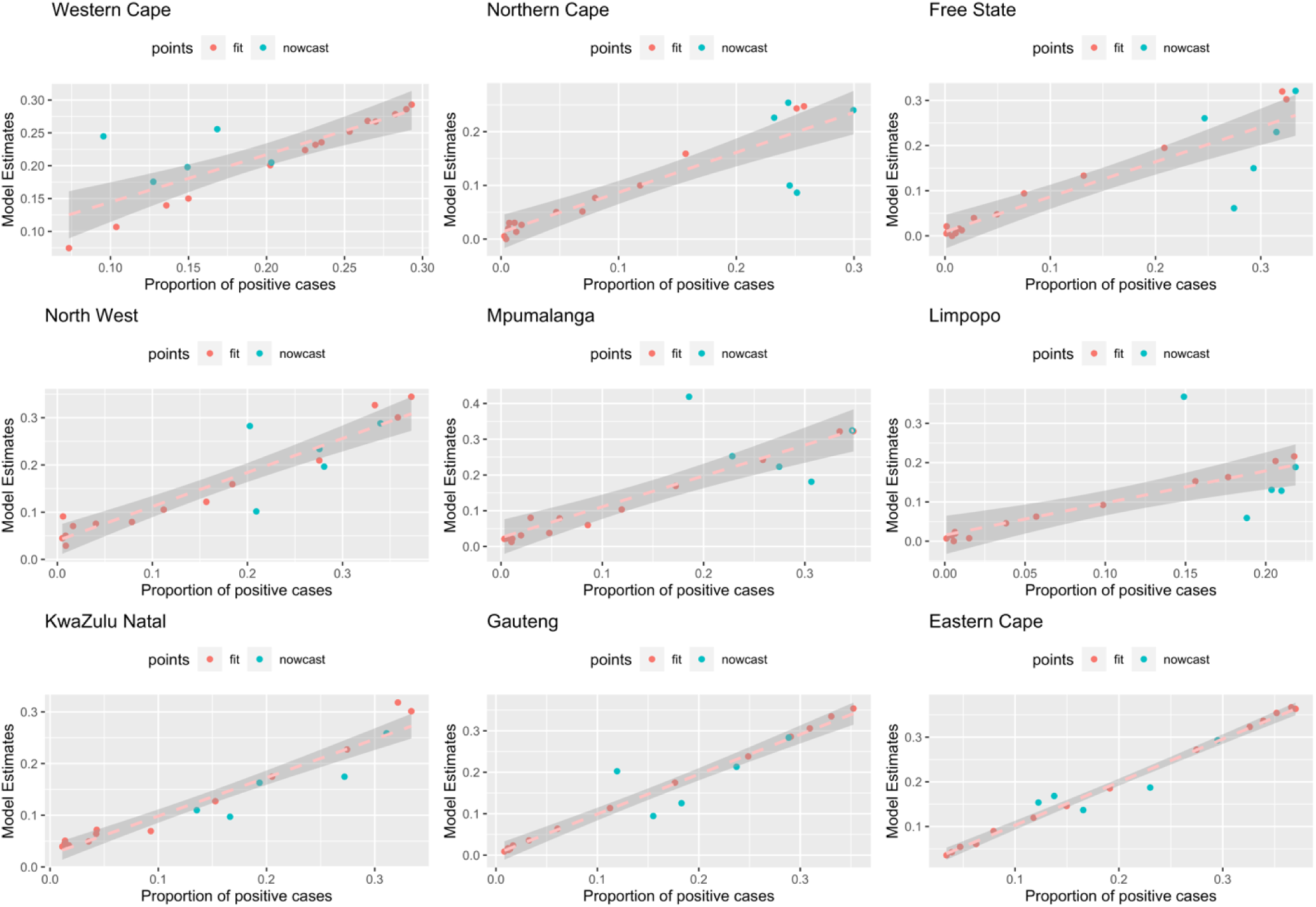
Association between model estimates and actual cases. Note that the model fit for certain provinces are much closer to the observed. We are using Google searches at time t to estimate cases at time t+1.

**SI Figure 2:**
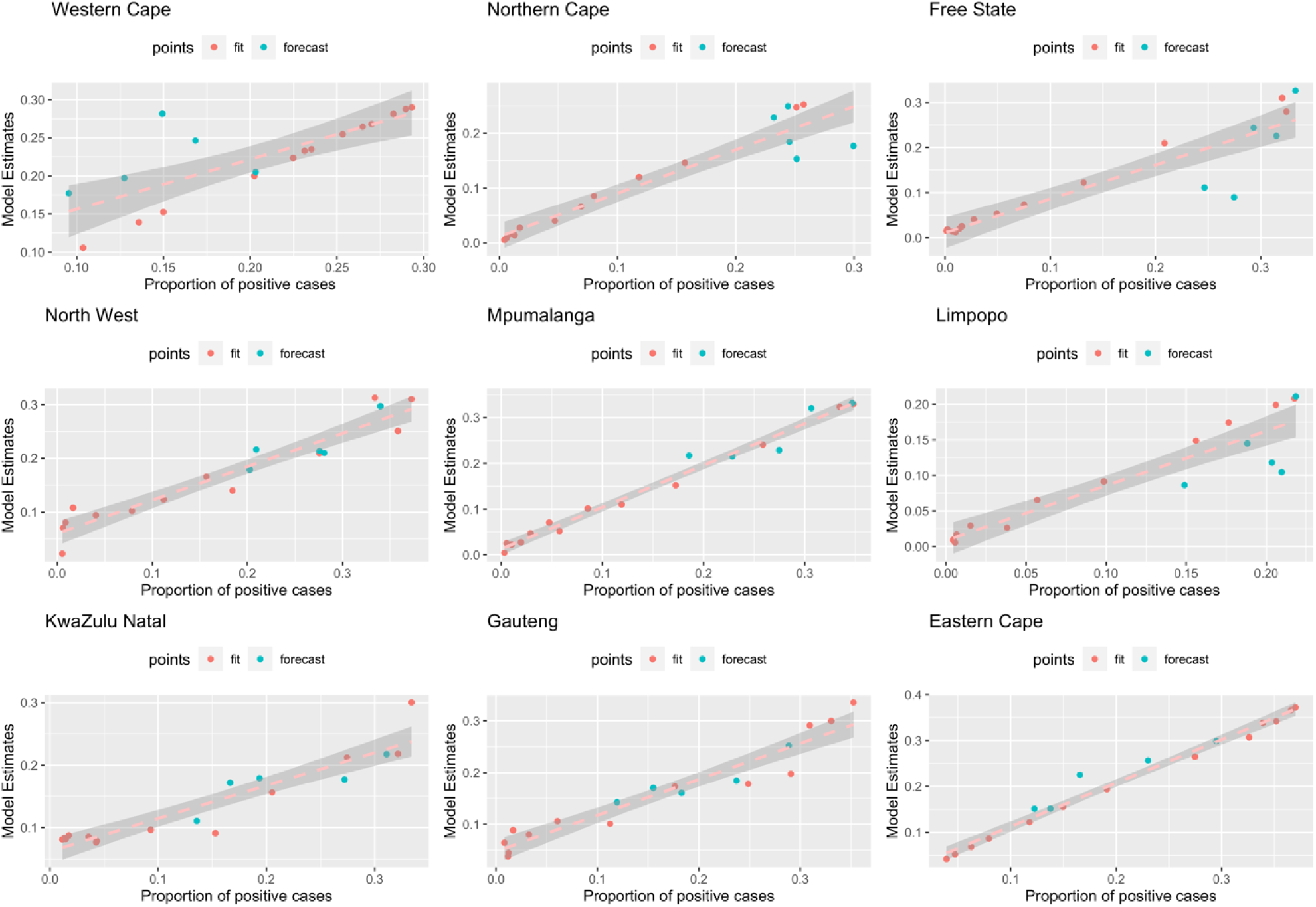
Association between model estimates and actual cases. Note that the model fit for certain provinces are much closer to the observed. We are using Google searches at time t to estimate cases at time t+1.

